# Defining a person-centered conceptual model to inform measurement of contraception’s effects on the menstrual cycle

**DOI:** 10.64898/2026.05.21.26353514

**Authors:** Amelia Mackenzie, Jenni Smit, Marija Miric, Alison Edelman, Mags Beksinska, Angely Catano, Stephanie Chung, Eunice Cuevas, Maddison Delacerda, Marci Messerle Forbes, Emily Hoppes, Leah Ingeno, Laura Jacobson, Mbali Khomo, Elena Lebetkin, Thandiwe Majola, Melissa Matos, Mbali Mavundla, Sarah McCaffrey, Alicia Mendez, Massiel Mendez, Nothando Mhlaba, Nzwakie Mosery, Lungelo Ndlovu, Bongeka Qiya, Kayla Stankevitz, Alexis Sullivan, Bongiwe Zulu

## Abstract

**Objective:** To address the need for improved measurement of the ways contraception impacts the baseline menstrual cycle (i.e., contraceptive-induced menstrual changes; CIMCs) by assembling an interdisciplinary, global research collective to rigorously develop a person-centered measure for CIMCs in multiple languages. As the first step, this paper reports on our conceptual model development, which is the foundation for ongoing measure development.

**Study design:** We conducted 18 focus groups with 106 people experiencing CIMCs while using hormonal or intrauterine contraception in Durban, South Africa, Santo Domingo, Dominican Republic, and Portland Oregon, United States. We used a virtual affinity mapping approach to analyze qualitative data, which was the basis of our conceptual model along with relevant theory and related models in the literature.

**Results:** The conceptual model of experiences with CIMCs depicts the baseline menstrual cycle, including CIMCs and conceptually-linked effects and the impacts and perceptions of those CIMCs. We found key domains of changes in pain, bleeding volume, bleeding patterns, and characteristics of blood.

**Conclusion:** Our CIMC conceptual model will inform development of a measure with evidence of validation across three language and global contexts. Adoption of a person-centered, standardized CIMC measurement across trials will improve knowledge and decision-making between methods.

**Implications:** Defining an evidenced-based conceptual model for CIMCs expands our understanding of how contraceptive users experience and understand menstruation and CIMCs. We can use this model to improve and standardize measurement of CIMCs.

## 1. INTRODUCTION

Changes in the baseline menstrual cycle while using hormonal or intrauterine contraception can influence sexual and reproductive health, daily lives, and overall wellbeing in both positive and negative ways.^1,2^ Referred to collectively as contraceptive-induced menstrual changes (CIMCs), this constellation of side effects and non-contraceptive benefits has been defined as: (a) changes in bleeding duration, frequency, volume, and predictability; (b) changes in characteristics of blood and other uterocervical fluid; (c) changes in uterine cramping and pain; (d) changes in other symptoms from the baseline menstrual cycle; (e) changes in experiences of menstrual and gynecologic disorders and symptoms (e.g., endometriosis, fibroids, adenomyosis); (f) and short-term changes to these factors after discontinuation.^1^

Recent efforts have aimed at improving how CIMCs are measured, including a systematic review of measurement of changes to the menstrual cycle across disciplines and expert consensus recommendations on measuring CIMCs in contraceptive clinical trials. ^3,4^ Findings from this work identify the need for a standardized, person-centered approach to CIMC measurement and call for the development of a novel CIMC measure with evidence of validation to support widespread use. The first step in developing a measure, and establishing that evidence base for the measures’ validity, is defining a person-centered conceptual model that diagrams conceptual domains of people’s experiences and the domain interrelationships to inform what should be measured.^5^ As no such model exists for CIMCs, we sought to define one as the foundation for our measure development.

## 2. METHODS

### 2.1. Data Collection

We conducted focus group discussions (FGDs) in Durban, South Africa, Santo Domingo, Dominican Republic, and Portland, Oregon US with purposively sampled adults of reproductive age (i.e., 18-49 years old) who reported experiencing CIMCs with current or recent (i.e., within three months) hormonal or intrauterine contraceptive use recruited from clinic-based and community-based settings. We used engaging approaches in FGDs, including: (a) an ideation card activity to guide participants in visualizing the CIMCs they experienced and the impacts on health, wellbeing, and daily lives, (b) an abbreviated body mapping exercise to explore menstrual pain and its locations, characteristics, and impacts, and (c) a “questerview” approach to elicit early interpretive feedback on common menstrual-cycle related questions. Facilitators conducted audio-recorded FGDs in isiZulu in Durban, Spanish in Santo Domingo, and English in Portland. Study teams used recordings, notes taken in each FGD, and, in some cases, transcription services to generate verbatim transcripts.

Ethics committees reviewed and approved the study protocol at the University of Witwatersrand’s Human Research Ethics Committee in South Africa, Two Oceans in Health’s internationally accredited independent IRB (IRB FWA00031272) and Consejo Nacional de Bioética en Salud in the Dominican Republic, and the Oregon Health & Science University’s IRB and FHI 360’s Protection of Human Subjects Committee in the United States. All study participants provided informed consent before data were collected and received compensation for their time and refreshments during FGDs.

### 2.2. Data Analysis

We used an adapted virtual affinity mapping approach to analyze FGD data. Traditional affinity mapping is a rapid, team-oriented methodology that can be used for qualitative analysis to collaboratively organize large amounts of data into themes, identify connections between themes, and incorporate insights from multiple team members with different levels of analysis experience.^6,7^ Our analysis methodology involved four steps following data collection, which we detail in Figure 1: (1) data organization; (2) individual preparatory work; (3) four virtual mapping sessions with all members of each partner study team; and (4) consolidation of mapping sessions into a single affinity map. We report more detail on our analysis approach in a separate paper.^8^

**Figure 1:**
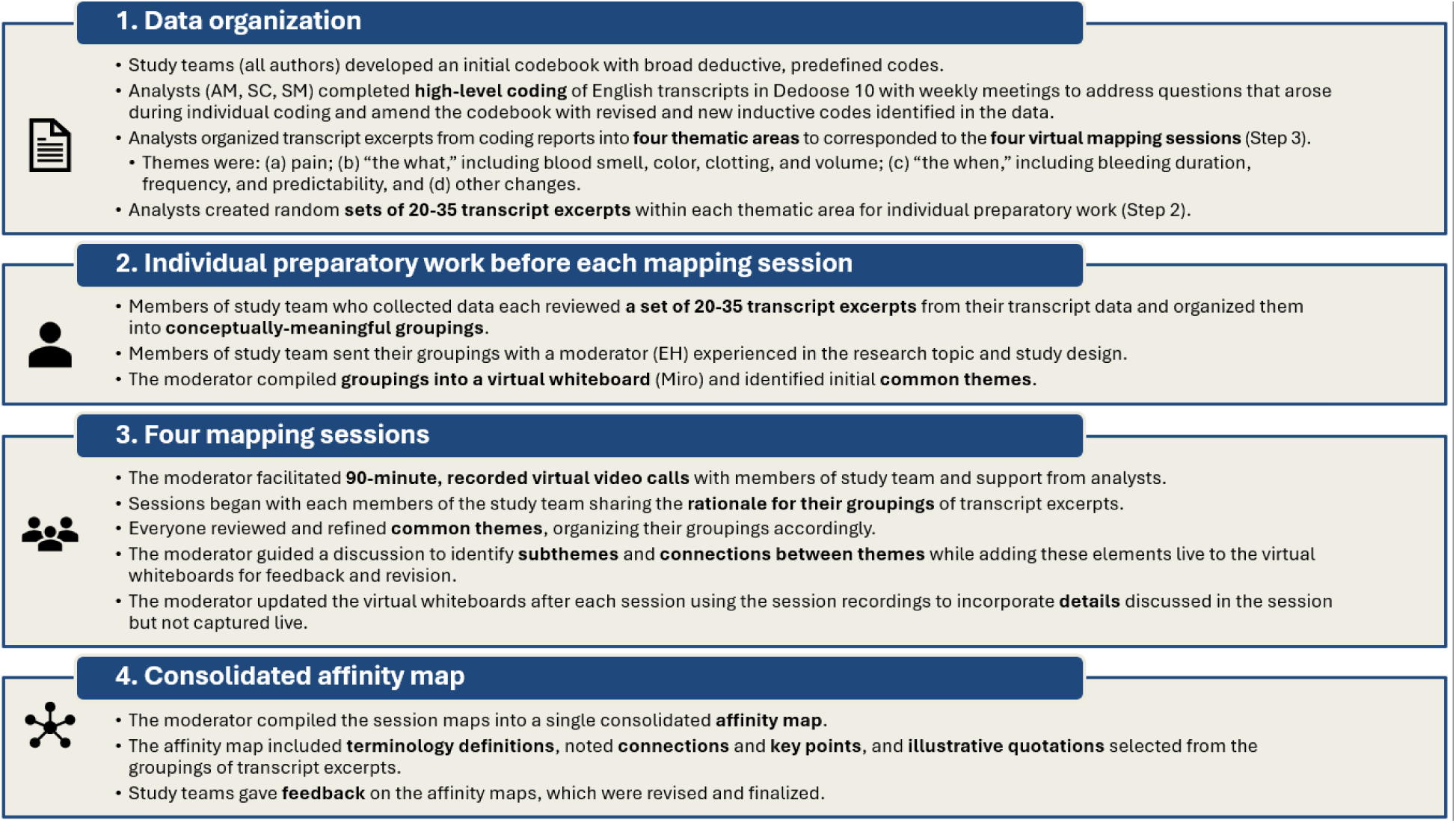
Virtual affinity mapping methodology.

### 2.3. Conceptual Model Development

Beginning with the consolidated affinity map, we incorporated elements of: (a) a measurement framework developed by the Global CIMC Task Force;^9^ (b) relevant theory; (c) a review of high-quality conceptual models from our recent systematic review of menstrual changes measurement;^3^ (d) related sexual and reproductive health models; and (e) conceptual models from other, related measures (Figure 2).

**Figure 2:**
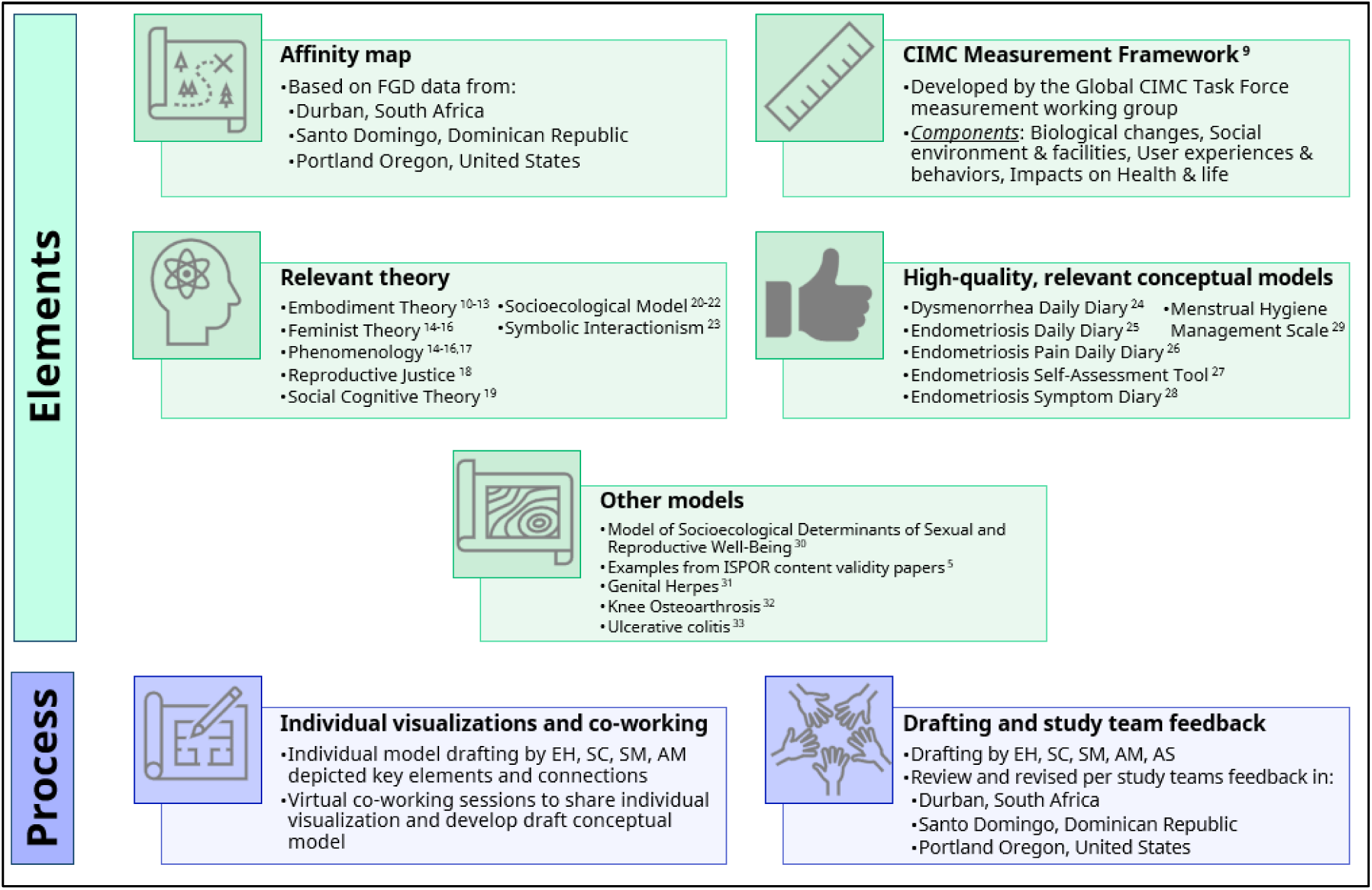
Development approach for the conceptual model.

## 3. RESULTS

### 3.1. Data collection and sample

We conducted 18 FGDs with 106 participants, four to seven per FGD: six in Durban, South Africa (n=39 participants); six in Santo Domingo, Dominican Republic (n=35); and six in Portland, US (n=32). Data collection took place between November 2024 and September 2025, and FGDs lasted an average of 84 minutes. We present participants characteristics in Table 1.

**Table 1:** Focus group discussion (FGD) participant characteristics by location in South Africa, Dominican Republic, and United States, 2024-2025.

|  | Durban | Santo Domingo | Portland | Total |
| --- | --- | --- | --- | --- |
| **Number of FGDs (participants)** | **6 (39)** | **6 (35)** | **6 (32)** | **18 (106)** |
| **Age** |  |  |  |  |
| Mean (SD) | 28 (6.4) | 30 (7.9) | 34 (8.1) | 31 (7.8) |
| Range | 18-44 | 18-45 | 20-49 | 18-49 |
| **Highest completed education (%)<sup>\*</sup>** |  |  |  |  |
| Primary | 0 | 6 | 0 | 2 |
| Secondary | 62 | 46 | 38 | 49 |
| Tertiary/University | 38 | 49 | 63 | 49 |
| **Partner (%)<sup>\*,\*\*</sup>** |  |  |  |  |
| None | 3 | 34 | 16 | 17 |
| Living apart | 85 | 90 | 31 | 43 |
| Cohabiting | 13 | 66 | 47 | 38 |
| **Contraceptive method (%) \*** |  |  |  |  |
| Pills | 5 | 40 | 34 | 25 |
| Injectables | 54 | 37 | 9 | 35 |
| Implants | 31 | 11 | 25 | 23 |
| Hormonal IUD | 3 | 3 | 19 | 8 |
| Copper IUD | 8 | 9 | 6 | 8 |
| Other | 0 | 0 | 6 | 2 |

\*Due to rounding, percentage totals do not all add up to 100%; \*\*2 participants did not report their partner status in Portland; “living apart” and “cohabiting” categories include married participants.

### 3.2. Conceptual model

We present a simplified version of our model in Figure 3, with the full model in Appendix A, including a broader map of the contraceptive experience within which our conceptual model is situated (see Appendix B). The model contains three components, with arrows indicating connections between and within components: (1) **changes in the baseline menstrual cycle**, including CIMCs and conceptually-linked effects; (2) **impacts of those changes**, organized by practical impacts, mental, emotional, and social impacts, and distal impacts of CIMCs; and (3) **perceptions of those changes**, including information about and perceptions of menstruation and contraception and the understanding of “normal” menstruation. For each component, we include illustrative quotations from our data in Table 2.

**Figure 3:**
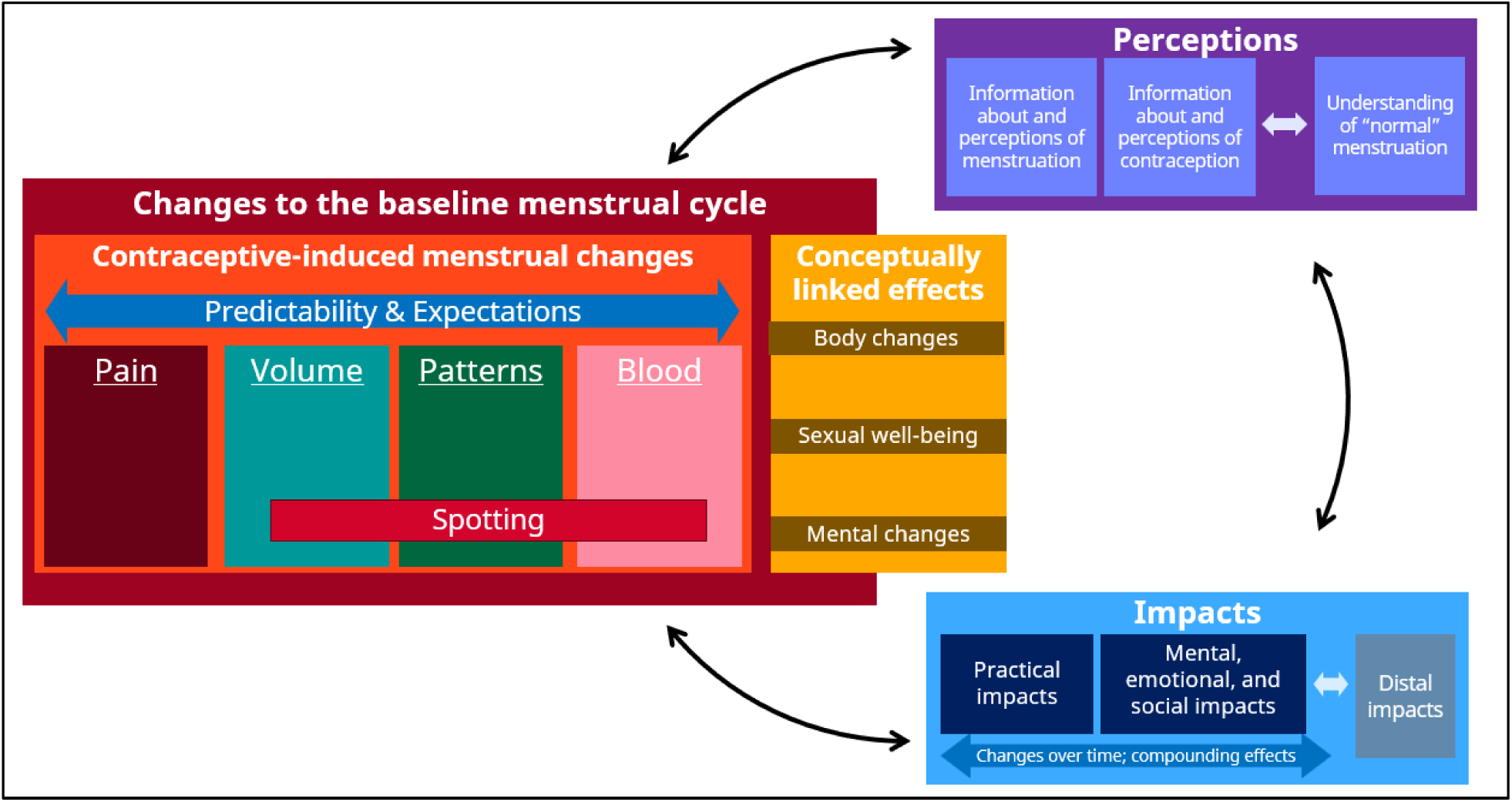
A simplified conceptual model of user experiences with contraceptive-induced menstrual changes (CIMCs).

**Table 2:**
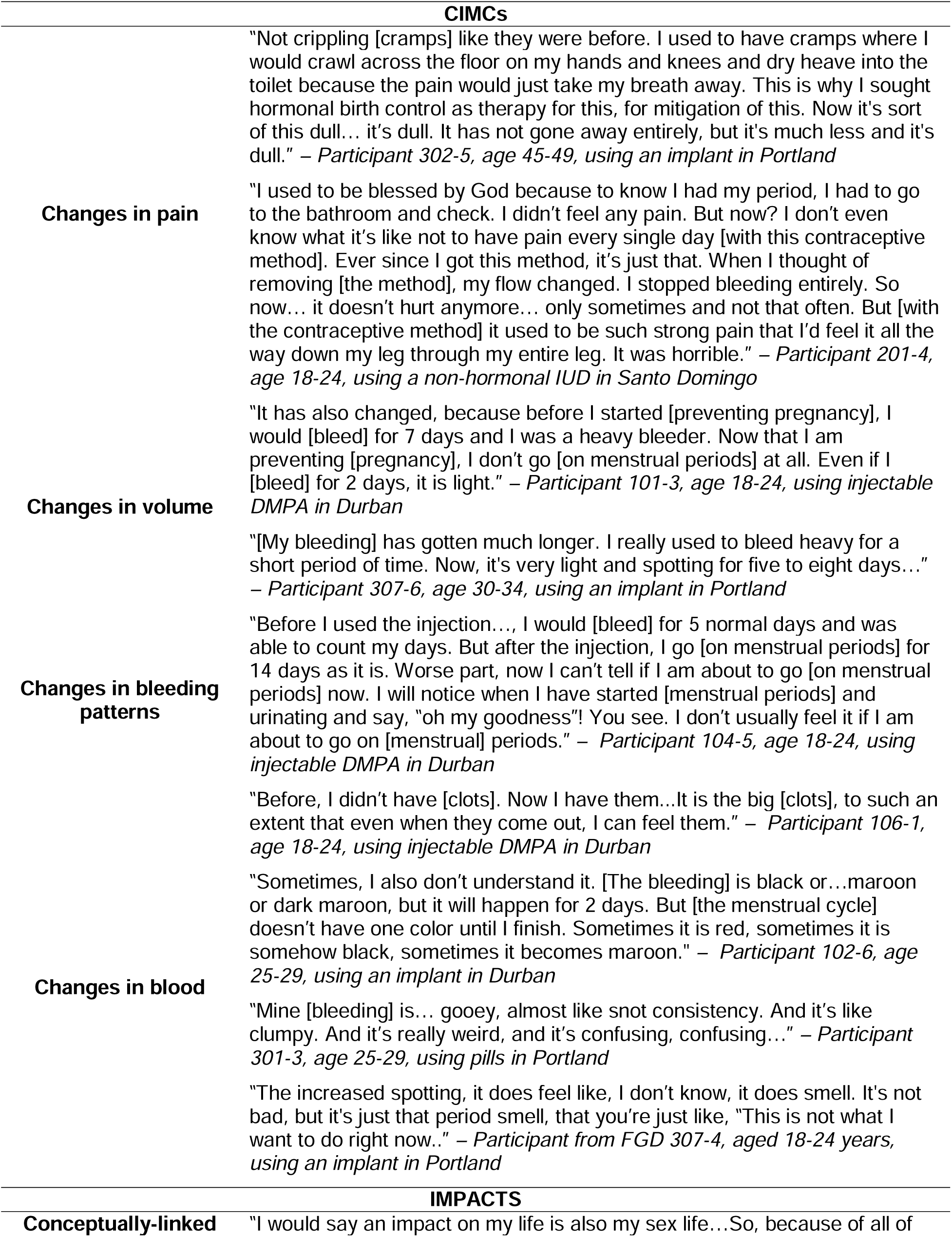
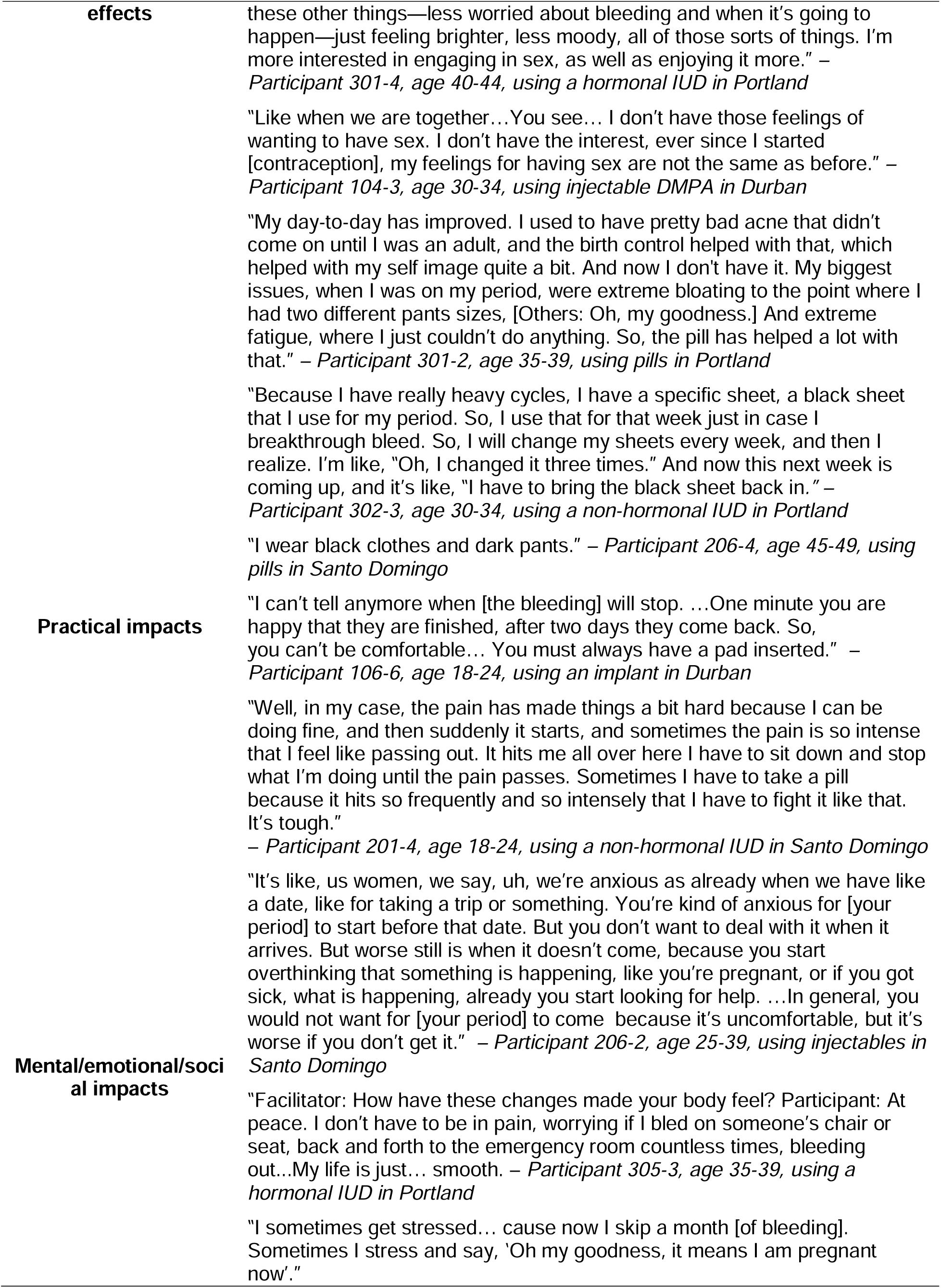
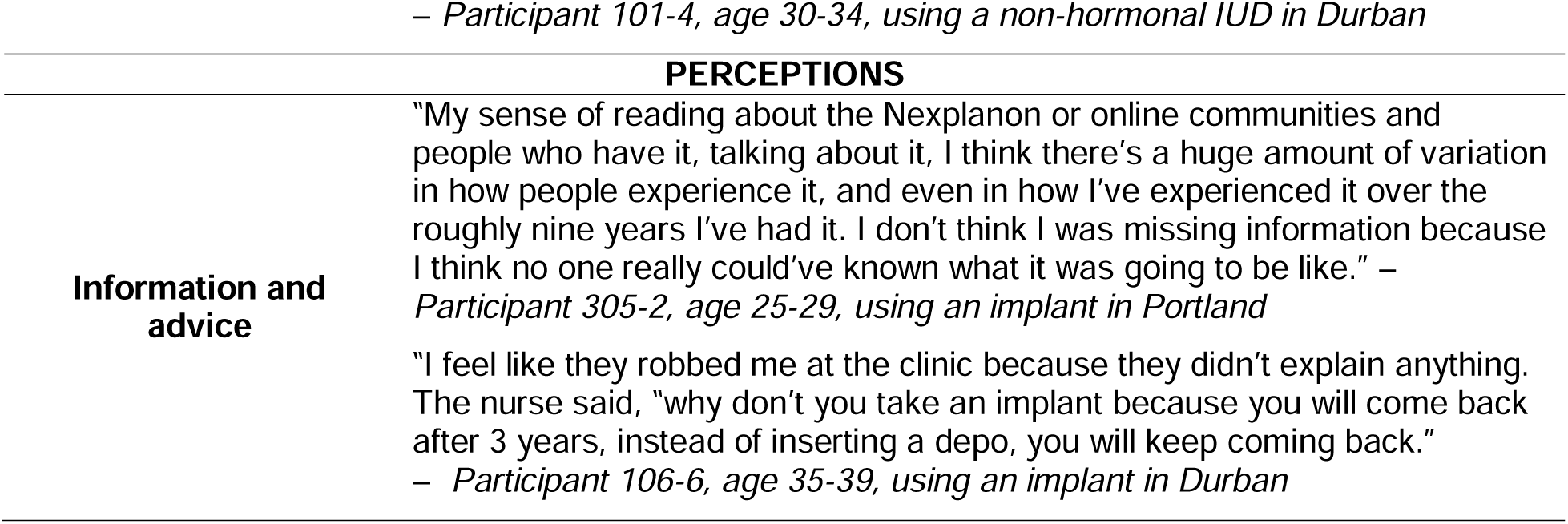
Illustrative quotations from FGDs for the three components of the conceptual model.

### 3.3. Model component: Changes in the baseline menstrual cycle

Details on the first component of the model are in Figure 4, depicting its two elements: CIMCs and conceptually linked side effects and non-contraceptive benefits. We identified four main domains of CIMCs: (1) changes in pain; (2) changes in bleeding volume; (3) changes in bleeding patterns; and (4) changes in blood characteristics, which we detail in the sections below. Conceptually linked effects are other changes that users consider being related to their menstruation or menstrual cycle. These are categorized by *body changes*, which can include changes in sleep, gastrointestinal aspects, skin, weight, inflammation, and more, and *mental changes*, including mood, psychosocial (e.g., a desire for isolation or feeling demotivated at work), and sensory changes. We call out *sexual well-being* separately, including sexual experience, libido, and non-menstrual bleeding, but these overlap with both body and mental changes. The full conceptual model provides further definitions, descriptors, and examples of each element and domain (See Appendix A).

**Figure 4:**
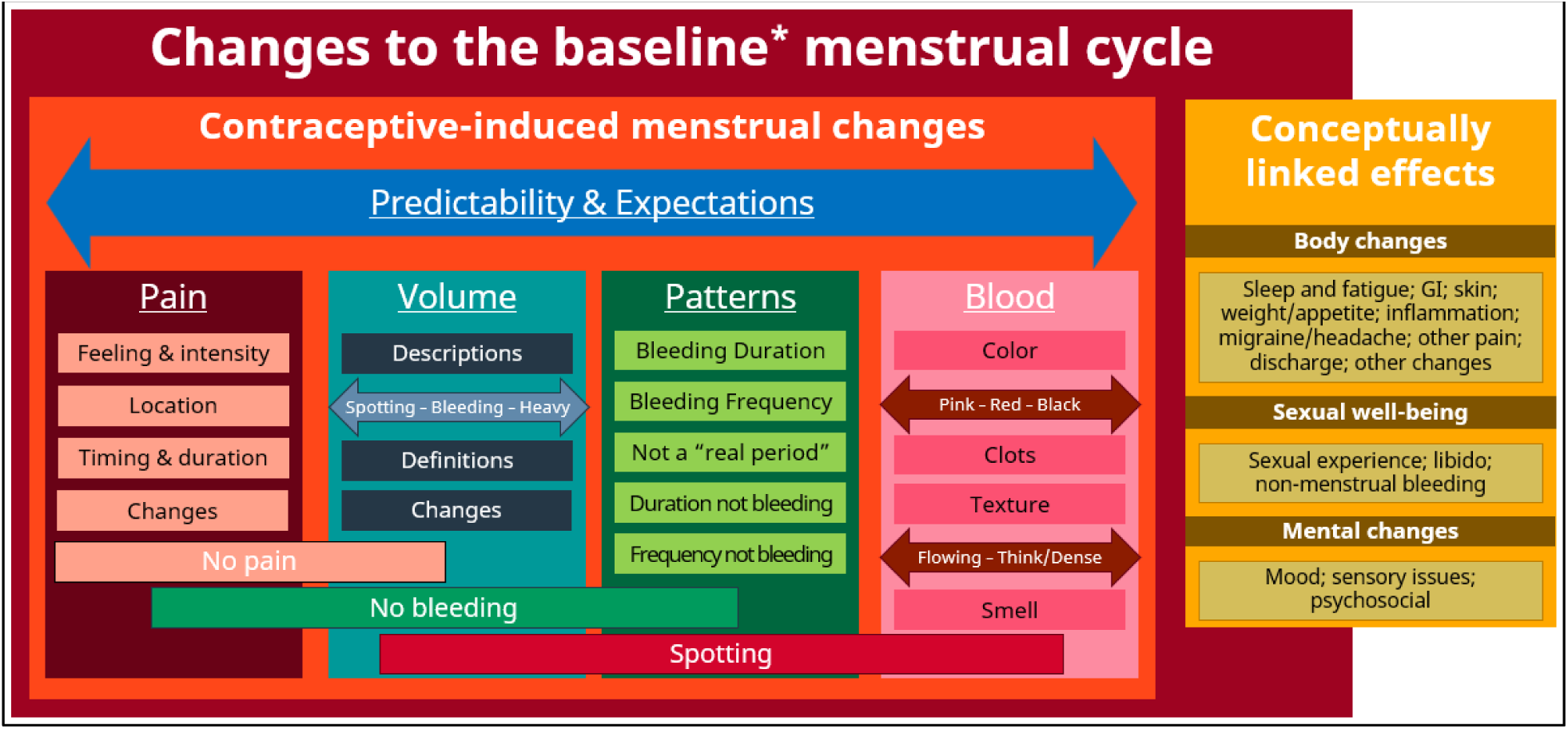
Changes in the baseline menstrual cycle: a component of the simplified conceptual model of user experiences with contraceptive-induced menstrual changes (CIMCs). *We use the term “baseline menstrual cycle” in the model to refer to the non-contracepting menstrual cycle to recognize the endometrial mechanisms causing bleeding with hormonal contraception are different from menstruation, and there’s no menstrual cycle. However, people using hormonal contraception often perceive the bleeding they experience while contracepting as part of their menstrual cycle, even if they are not experiencing physiological menstruation.

#### 3.3.1. Domain: Changes in pain

Pain includes a variety of characteristics: *feeling and intensity*, *location*, *timing and duration*, and *changes in pain*. *Absence of pain* is also included in this domain. *Feeling and intensity* is defined as the experience, sensation, amount and/or intensity of pain felt in the body and can be described on a spectrum from no pain to intense pain. *Location* is the area of the body where pain is felt/experienced, while *timing and duration* includes descriptions of the length of time pain occurs, often described alongside when it occurs. *Changes in pain* are the variations in the degree, amount, location or length of pain.

#### 3.3.2. Domain: Changes in volume

Volume is characterized by *definitions and descriptors* of how much blood comes out, and *changes in volume*, or variations in how much blood comes out. Volume is often described or defined along a spectrum from spotting to very heavy bleeding, by the type of menstrual product, or by how frequently menstrual products need to be changed. *No period/bleeding*, as well as *no pain*, is also included in this domain and is defined by bleeding completely stopping for an extended amount of time. *Spotting*, or very small amounts of blood that is often unpredictable, is also included in this domain.

#### 3.3.3. Domain: Changes in Bleeding Patterns

Changes in patterns is organized into two sections: *time with bleeding* and *time without bleeding*. Within *time bleeding*, the following characteristics are identified: *duration*, or the number of days of bleeding, *frequency of bleeding*, or how often bleeding occurs, and *not a ‘real period’*, or uncertainty around whether what is being experienced is a menstrual period or not.

Within *time without bleeding*, *duration* and *frequency of time* without bleeding, or how often time without bleeding occurs, are described. *No period/bleeding* and *Spotting* extends to this domain as well.

#### 3.3.4. Domain: Changes in blood

Changes in blood characteristics features multiple characterstics: *blood color*, *blood texture*, *clots*, and *blood smell*. *Blood color* is defined as the shade or pigment of the blood when it comes out of the body. *Blood texture* is defined by the descriptive words used to explain the texture of blood and can be described on a spectrum from light, liquid and flowy to dense, compact and thick. *Clots* include four factors: consistency, color, size and feeling. Finally, *blood smell* is the odor of the blood when it comes out of the body. As with changes in volume and patterns, s*potting* extends to this domain.

#### 3.3.5. Connections between CIMC domains

Some characteristics are relevant across domains. *Expectations* and *predictability* span the four domains above. We define *expectations* as what a person believes will or should happen based on wider context and perceptions of and experiences with menstruation and contraception. This includes the idea of normalcy and what someone believes a “normal” menstrual cycle or bleeding pattern looks like. *Expectations* play a role in changes in pain, volume, patterns, and blood. We define *predictability* as whether any CIMC is anticipated or not and/or regular or not. This is often informed by menstrual and contraceptive history and ongoing menstrual and contraceptive experiences. *Predictability* plays a role in changes in pain, volume, patterns, and blood. As noted, the component of *no bleeding* applies to the volume and patterns domains. Similarly, *spotting* spans across volume, patterns and blood.

Other connection points are displayed only in the full conceptual model with grey circles, representing areas where two or more domains, or characteristics within a domain, are connected (see Appendix A). For example, we found connections beetween changes in pain and changes in volume, describing how changes in pain often happen in conjunction with other changes, like changes in volume and changes in blood consistency. Similarly, we found a connection between changes in volume and changes in bleeding patterns, explaining duration, frequency, and amount of bleeding are often discussed together and are not always distinguishable. Within changes in blood, we found a connection between blood color, texture, timing, and normality. Importantly, a connection point for the overall CIMCs section clarifies that *all* changes are linked and were discussed in overlapping and even interchangeable ways.

### 3.4. Model component: Impacts

The impacts of CIMCs are an important consideration for users. Impacts can be from more than one CIMC or changes in CIMCs over time, and they can have compounding effects. Impacts are grouped into (a) practical impacts; (b) mental, emotional, and social impacts, and (c) distal impacts (Figure 5; Appendix A).

**Figure 5:**
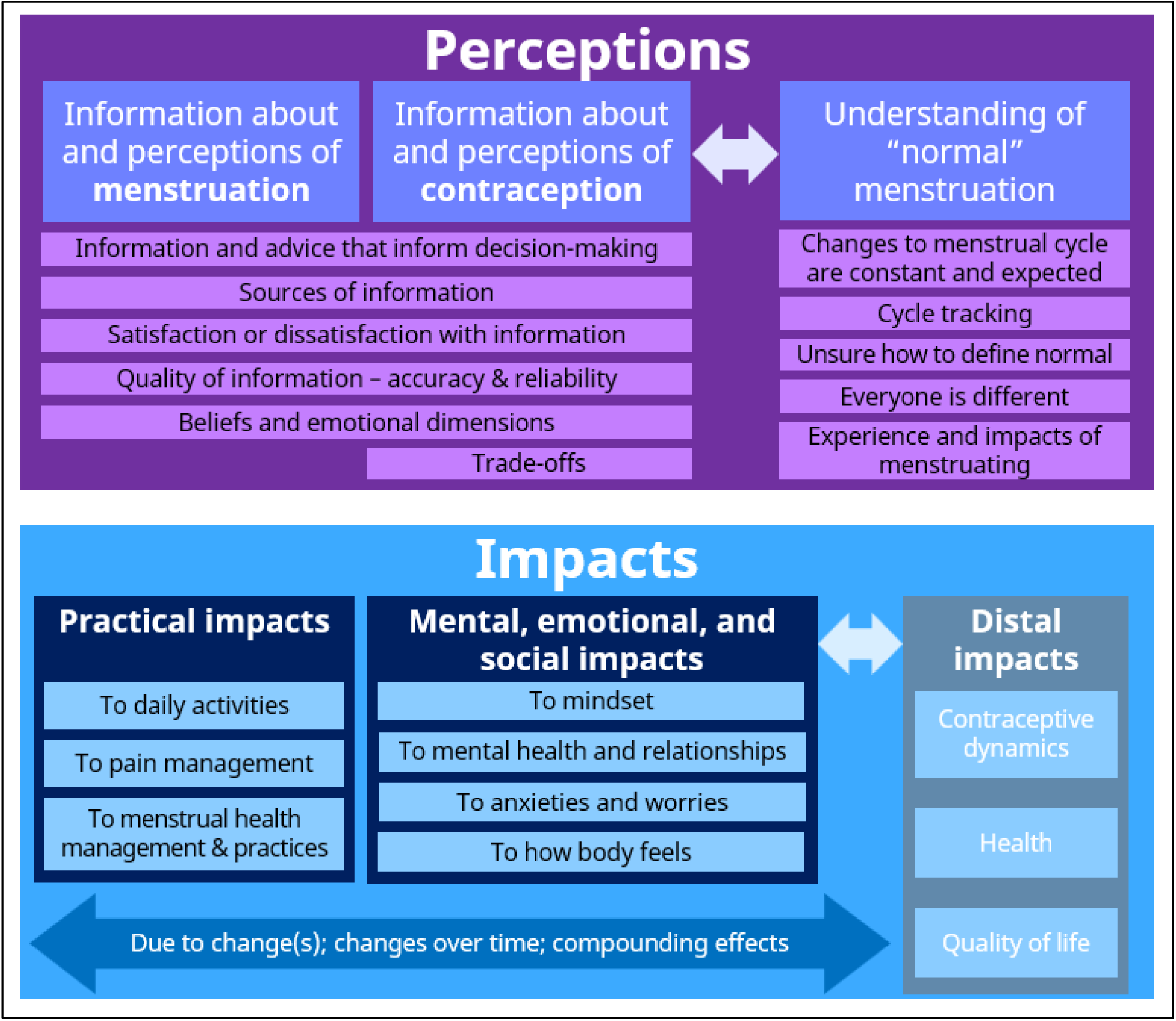
Impact and perceptions of contraceptive-induced menstrual changes (CIMCs): Components of the simplified conceptual model of user experiences with CIMCs.

In terms of practical impacts, *impacts to daily activities*, or the effects on a person’s ability to carry out routine tasks and responsibilities of their lives, applies to all four domains of CIMCs (Figure 4). *Impacts to pain management/relief*, or affects on the methods used to manage or reduce menstrual pain, falls under the changes in pain domain. Finally, *impacts to menstrual product use* is relevant to the changes in volume, changes in patterns, and changes in blood domains.

Mental, emotional, and social impacts of the changes in pain and changes in volume domains include *impacts to mindset*, or changes or shifts in attitudes or approaches towards pain. *Impacts to mental health and relationships*, or changes or shifts in emotional, psychological, and social well-being, also applies to the changes in pain domain. *Impacts to anxieties/worries*, or changes or shifts in concern or fear related to health, safety, or personal well-being, falls under the changes in volume, patterns and blood domains. Finally, *impacts to how body feels*, or changes or shifts in physical sensations and perceptions experienced, is relevant to the changes in patterns and changes in blood domains.

Distal impacts of CIMCs include *impacts to contraceptive dynamics*, such as contraceptive non-use, choice, satisfaction, and more, *impacts to health*, including unintended pregnancy, symptom management for menstrual and gynecological disorders, and management of anemia or iron deficiency, and *impacts to quality of life*, such as psychosocial well-being, sexual well-being, gender affirmation and more.

Finally, there are some connection points, notated in grey circles in Appendix A, throughout the impacts component of the model. For example, among mental, emotional, and social impacts, we found a connection between loss of libido and bleeding, often combining to cause relationship impacts. Here also, we found a connection within the changes in pain and changes in bleeding patters, where the changes in pain were often more emotionally charged than the changes in bleeding patterns.

### 3.5. Model component: Perceptions

Perceptions of menstruation and contraception emerged as an important component of how users conceptualize CIMCs (Figure 5; Appendix A). Experiences with CIMCs are shaped not only by the characteristics of the changes themselves, but also by how individuals interpret and make sense of those changes through informational, social, and cultural frameworks. In this way, perceptions can mediate between CIMCs and their impacts.

First, people spoke about *information about menstruation and perceptions of menstruation*. This can include the following: *Information and advice*, which is the knowledge, insights, and guidance that inform decisions about mensruation, menstrual product use, or seeking care for menstrual disorders. *Source of information* is the people and channels that provide information about menstruation, reproductive health, and contraceptive-induced menstrual changes that shape someone’s expectations of future experiences. *Satisfaction with information* is whether health information about menstruation was satisfactory or lacking, while *quality of information* is the accuracy and reliability of information about menstruation. *Menstrual beliefs* are the perspectives and conceptions one holds regarding menstruation. Finally, *belives and emotional dimensions* are how people feel when learning or talking about menstruation.

Secondly, *information about contraception and perceptions of contraception* included many of the same components, including *information and advice*, *sources of information*, *satisfaction of information*, *quality of information*, *contraceptive beliefs*, and *emotional dimensions*. *Trade-offs* was added, which is the idea that users weigh the benefits and costs of contraceptive use as they form opinions and make decisions about methods and the CIMCs associated with them.

Finally, *understanding of “normal” menstruation* captures five categories. *Change as constant* reflects the view that changes to the menstrual cycle are constant and expected. *Cycle tracking* is monitoring and recording various aspects of the menstrual cycle using a variety of methods. *Undefined normal* is confusion about what is defined as normal for both menstruation and CIMCs. *Everyone is different* captures the idea that everyone experiences CIMCs and other side effects differently, while *the menstrual experience* is the idea that people who do not menstruate may not understand its importance in the lives of those who do.

## 4. DISCUSSION

We define a comprehensive conceptual model of CIMC experiences among people using contraception based on data from interactive FGDs across three different global settings as well as existing models, frameworks, and relevant theory. With this comprehensive approach, we ensured the model was grounded in findings from our data while also highlighting existing evidence within a strong theoretical base. Importantly, our model also moves beyond a descriptive list of CIMCs to acknowledge the relevance of how people perceive, interpret, and experience these changes and the connections between them.

An evidence-informed, person-centered conceptual model like this can be the foundation for improving measurement, both broadly but also in the context of clinical trials. The use of patient-reported outcome (PRO) measures in clinical trials is recognized as important tools by regulators, clinicians, and patients for outcomes reported directly by trial participants about the status of their health condition, including unobservable effects known only to participants (e.g., pain) and participants’ perspectives on those effects and their impacts.^34,35^ Therefore, our approach here is grounded in FDA guidance and PRO development best practices.^35,,36^ In addition, as far as we are aware, our effort is the first PRO development for clinical trial use that follows recommended practices for establishing transferability and instrument translation via simultaneous development in three global regions and in three languages (i.e., isiZulu and English in South Africa, Spanish in the Dominican Republic, and English in the United States), permitting cross-cultural comparison of potentially diverse menstrual and contraceptive experiences.^37,38^ This type of rigorous approach is largely unique in the field of contraceptive development. Work on other contraceptive side effects and non-contraceptive benefits, including sexual wellness and pleasure, can apply our rigorous approach, including careful attention to perceptions and impacts of the changes experienced with contraception. This comprehensive view and understanding of CIMCs can also inform future research into some of the domains identified in our model that have not been well-studied (e.g., changes in characteristics of blood and other uterocervical fluid) and are not yet fully understood. Measurement of CIMCs and other side effects in contraceptive clinical trials using a standardized, person-centered measure with evidence of validity across contexts has the potential to improve product labeling for new contraceptive methods. It will also allow greater comparability across trials and between methods, which can inform clinical guidance and provider counseling, thus providing people using contraception with accurate and relevant information to make truly informed decisions about their health, wellbeing, and quality of life.

## Data Availability

All data produced are available online via Harvard Dataverse (https://doi.org/10.7910/DVN/OF8N4O)

## ACKNOWLEDGMENTS

The authors would like to recognize the additional contributions of Vivian Brache and Leila Cochón from Profamilia for their input on the study protocol and engagement on study calls. We also appreciate the members of the study’s Technical Advisory Group: Professors Kathryn Clancy of the University of Illinois, Urbana-Champaign, Elizabeth Raymond of the University of Washington, Funmi OlaOlorun of the University of Ibadan, and Carolina Sales Vieira of the University of São Paulo. We appreciate our partnerships with Kobo Toolbox and Biologics Consulting for additional technical guidance, especially Tino Kreutzer, Salomé Garnier, Ignacio Giménez Rebollo, Chrissy Roberts, Margaret Vernon, Samira Shirwa, as well as Lisa Soule.

The FHI 360 team thanks Rebecca Callahan, Kavita Nanda, Laneta Dorflinger, Douglas Taylor, and Neha Mehta for their technical guidance and *ad hoc* support of the study, and Madison Wojtkowiak for her support of the study start up. The OHSU team thanks Maggie McBride for her assistance in study recruitment. We thank Megan Christofield for her discussion on the affinity mapping methodology, which sparked the idea for our analysis approach of the FGD data. We also appreciate feedback from Rebecca Callahan on the manuscript.

## ACKNOWLEDGEMENT OF FUNDING

This work was supported by the Gates Foundation (INV-062479). The conclusions and opinions expressed in this work are those of the author(s) alone and shall not be attributed to the Foundation or FHI 360. Under the grant conditions of the Foundation, a Creative Commons Attribution 4.0 License has already been assigned to the Author Accepted Manuscript version that might arise from this submission.

## Notes

### Competing Interest Statement

AE has received travel reimbursement from ACOG, WHO, CDC, and honoraria from Gynuity for committee activities. AE receives royalties from Up to Date, Inc. Oregon Health & Science University receives research funding from OHSU Foundation, Gates, Merck, HRA Pharma, and NIH where AE is the principal investigator. All other authors have no conflicts of interest to declare.

### Funding Statement

This study was supported by the Gates Foundation (INV-062479). The conclusions and opinions expressed in this work are those of the author(s) alone and shall not be attributed to the Foundation or FHI 360. Under the grant conditions of the Foundation, a Creative Commons Attribution 4.0 License has already been assigned to the Author Accepted Manuscript version that might arise from this submission. Please note works submitted as a preprint have not undergone a peer review process.

### Author Declarations

The Human Research Ethics Committee of University of Witwatersrand gave ethical approval for this work. The IRB of Two Oceans in Health gave ethical approval for this work. The Consejo Nacional de Bioetica en Salud of the Dominican Republic gave ethical approval for this work. The IRB of Oregon Health & Science University gave ethical approval for this work. The Protection of Human Subjects Committee of FHI 360 gave ethical approval for this work.

