## Appendix A for "Defining a person-centered conceptual model to inform measurement of contraception’s effects on the menstrual cycle"

**Appendix A:** Full conceptual model User Experiences with Contraceptive-Induced Menstrual Changes (CIMCs)

Go to the following website to see the full model: https://miro.com/app/board/uXjVGjbEAeI=/?share_link_id=799097710708
