## Appendix B for "Defining a person-centered conceptual model to inform measurement of contraception’s effects on the menstrual cycle"

**Appendix B.** The contraceptive experience context for our conceptual model

While developing the conceptual model, we determined we first needed to define a broader contraceptive experience in which our conceptual model is situated. The contraceptive experience includes **three elements**: (a) the *larger social contex*t, which includes immediate social environments (e.g., family, friends, peers) and structural or systemic contexts (e.g., workplace, healthcare access, cultural norms, and policy); (b) the *individual context*, which includes individual identity, life stage, and how these relate to past menstrual and contraceptive experiences; and (c) the *menstrual experience*. We also defined **two states** within the contraceptive experience: (1) before (or between or after) contraceptive use, and (2) during contraceptive use, which are connected by the introduction of a contraceptive method. Our conceptual model of user experiences with CIMCs sits within the menstrual experience component during contraceptive use.

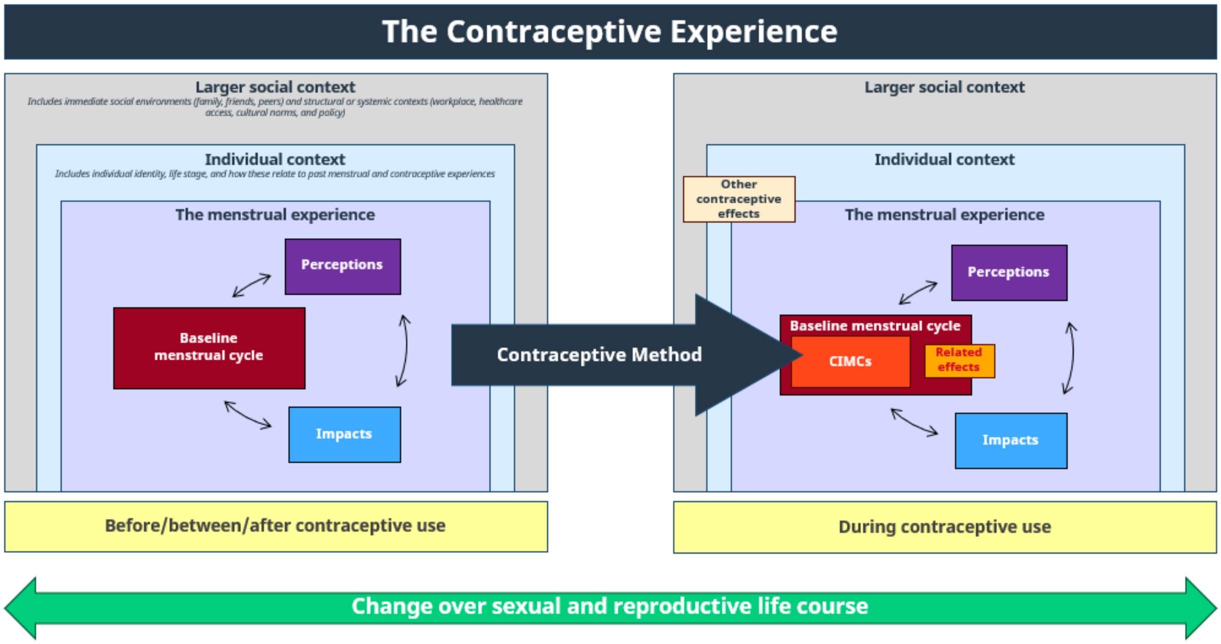
